# Detection of two CAL.20C SARS-CoV-2 variants in Monterrey metropolitan area in Northeast Mexico

**DOI:** 10.1101/2021.04.05.21251104

**Authors:** Kame A. Galán-Huerta, Ana M. Rivas-Estilla, Sonia A. Lozano-Sepúlveda, Natalia Martínez-Acuña, Daniel Arellanos-Soto, Roberto Montes-de-Oca-Luna, Consuelo Treviño-Garza, Manuel E. de-la-O-Cavazos, Javier Ramos-Jiménez

## Abstract

SARS-CoV-2 variants of concern (VOC) are a worldwide problem. CAL.20C is considered a VOC and its distribution should be monitored. We detected two CAL.20C variants in Monterrey, Mexico in patients who had no recent travel history. This indicates that this variant is now transmitted locally in Northeast Mexico.

---

Since September 2020, several SARS-CoV-2 variants of concern (VOC) have been emerged. Worldwide, B.1.1.7, B.1.351, and P.1 variants are monitored because they are more transmissible compared to other lineages. B.1.1.7 variant has been also suspected to be more lethal (*1*), and B.1.351 associated with lower vaccine efficacy (*2*). In October 2020, an increase in COVID-19 cases in southern California led to the identification of a new variant designated CAL.20C (20C/S:452R;/B.1.429) (*3*). Recently, the Centers for Disease Control and Prevention have classified variants 20C/S:452R (B.1.427 and B.1.429) as VOC (https://www.cdc.gov/coronavirus/2019-ncov/cases-updates/variant-surveillance/variant-info.html).

The Center for Investigation and Innovation in Medical Virology (CIIMV), in Monterrey, Mexico, has been screening SARS-CoV-2 positive samples with low Ct values looking for these variants in Northeast Mexico.

We analyzed 54 RNA samples positive for SARS-CoV-2 by RT-qPCR (*4*) with Ct values lower than 20. The samples were collected from November 14, 2020 – January 22, 2021, from Nuevo Leon’s Public Health State Laboratory. Whole genome sequencing was done with Artic Network’s amplicon sequencing (*5*) in MinION (Oxford Nanopore Technologies, Oxford, UK. Multiple sequence alignment was done in MAFFT online service (*6*). Maximum clade credibility tree was inferred using BEAST v1.10.4 (*7*) and used the Pango lineage nomenclature (*8*). Sequences generated in this report were deposited in GISAID with the following numbers: EPI_ISL_979342 and EPI_ISL_1091256. This work was reviewed and approved by the Ethics Committee from Facultad de Medicina y Hospital Universitario Universidad Autónoma de Nuevo León with the following registration number: BI20-0004.

We found 22 different SARS-CoV-2 lineages circulating in Nuevo Leon state. Lineages B.1.1.7, B.1.351, or P.1 were not detected. Nevertheless, the California lineages B.1.429 and B.1.427 were found in two residents of Nuevo Leon. The samples belonged to two females with no travel history in the previous week.

The first patient, where lineage B.1.429 was detected, had onset of symptoms the first days of January 2021. This patient was in her 20’s, with no comorbidities, and only reported cough and headache. The second patient, where lineage B.1.427 was detected, had onset of symptoms in mid-January 2021. This patient was in her 60’s, with no comorbidities, and reported cough, headache, odynophagia, myalgia, arthralgia and rhinorrhea. Both patients were treated ambulatory.

Both variants (NLE-UANL-005/2021 and NLE-UANL-025/2021) had the characteristic spike mutations L452R and W152C, orf1ab: D5584Y and N: T205I. Additionally, sample NLE-UANL-005/2021 had the orf1a I4205V mutation. Phylogenetic analyses place these two samples within viruses isolated in California.

Sample NLE-UANL-005/2021 (lineage B.1.429) grouped in a well-supported clade with samples isolated in California (Figure). The most recent common ancestor of this clade circulated in mid-December (95% CI November 28, 2020 – December 25, 2020). Because the patient denied recent travel, we can speculate that the lineage was introduced to the state and spread during the holidays.

**Figure.**
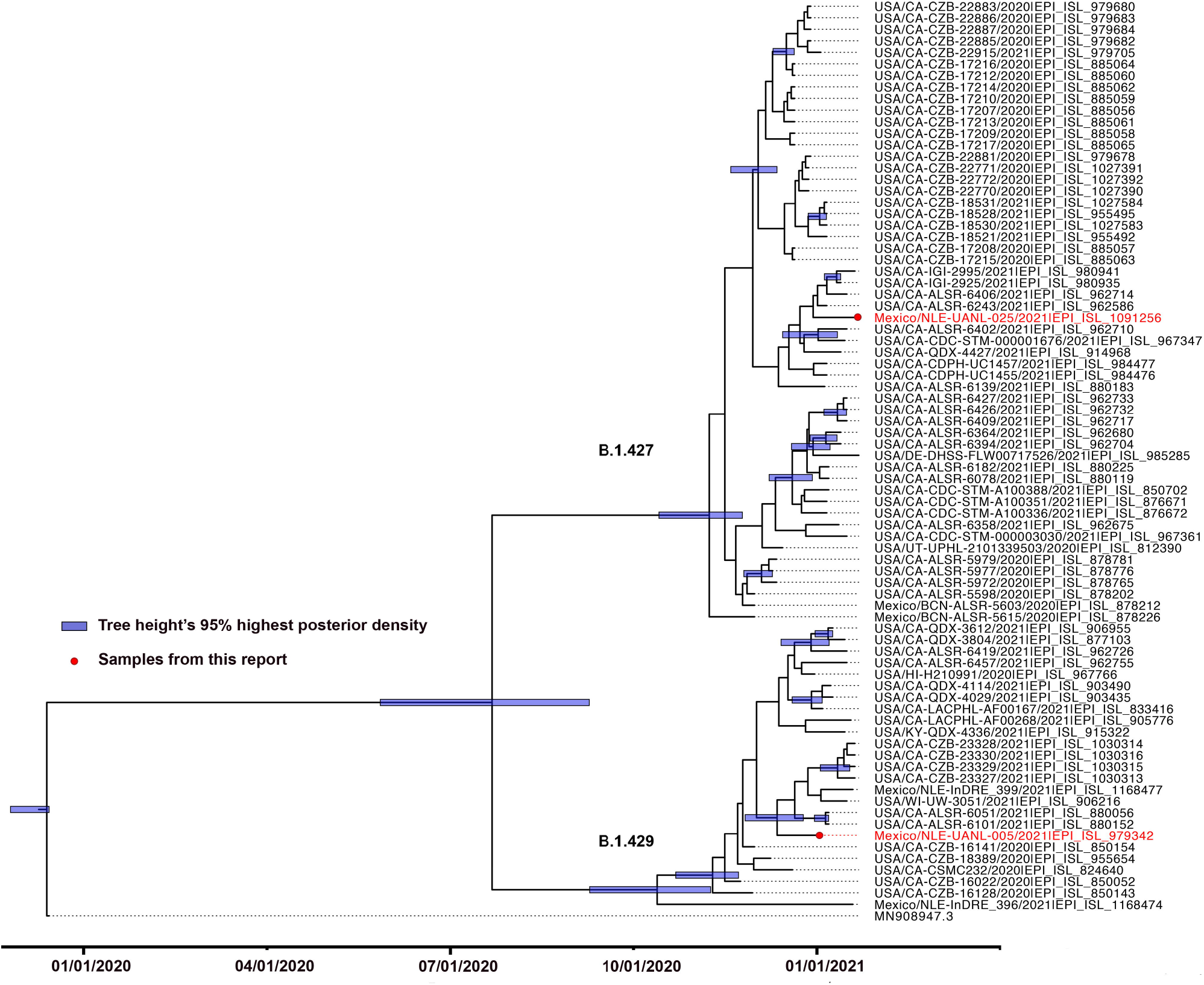
Phylogenetic tree showing the relationship of the sequenced viruses (n = 80) and the previously reported CAL.20C variants. The tree was inferred in BEAST v.1.10.4 using HKY+G substitution model, strict clock, coalescent exponential growth tree prior, and a 50 million chain length. Nodes with high posterior probability are indicated with rectangles. Samples generated in this report are indicted with a circle. Sequences used in this analysis were downloaded from GISAID. The authors of sequences used in this report are listed in the Appendix Table.

Sample NLE-UANL-025/2021 (lineage B.1.427) does not group in a well-supported clade. The most recent common ancestor of samples belonging to this lineage was estimated to appear the first days of November (95% CI October 14, 2020 – November 24, 2020).

Because of the potential epidemiological implications for a raise in case count with these variants, the health authorities in neighboring northeast Mexican states, as well as Texas and other southern US states should be aware of the presence, although limited by number, of these variants.

## Data Availability

The sequences generated were deposited in GISAID. https://www.gisaid.org/

## Acknowledgments

This work was funded by Consejo Nacional de Ciencia y Tecnología (CONACyT) with the following grant number 312328, awarded to Kame A. Galán-Huerta. The authors gratefully acknowledge the authors and submitting laboratories that generated genome data and shared it via GISAID. A full list acknowledging the authors submitting data used in this report can be found in Appendix Table.

## References

1. Challen R, Brooks-Pollock E, Read JM, Dyson L, Tsaneva-Atanasova K, Danon L. Risk of mortality in patients infected with SARS-CoV-2 variant of concern 202012/1: matched cohort study. BMJ. 2021 Mar 9;372:579. doi: 10.1136/bmj.n579. PubMed PMID: 33687922; PubMed Central PMCID: PMC7941603.

2. Madhi SA, Baillie V, Cutland CL, Voysey M, Koen AL, Fairlie L, et al.Efficacy of the ChAdOx1 nCoV-19 Covid-19 Vaccine against the B.1.351 Variant. N Engl J Med. 2021 Mar 16. doi: 10.1056/NEJMoa2102214. Epub ahead of print. PubMed PMID: 33725432.

3. Zhang W, Davis BD, Chen SS, Sincuir Martinez JM, Plummer JT, Vail E. Emergence of a Novel SARS-CoV-2 Variant in Southern California. JAMA. 2021 Feb 11:e211612. doi: 10.1001/jama.2021.1612. Epub ahead of print. PubMed PMID: 33571356; PubMed Central PMCID: PMC7879386.

4. Corman VM, Landt O, Kaiser M, Molenkamp R, Meijer A, Chu DK, et al. Detection of 2019 novel coronavirus (2019-nCoV) by real-time RT-PCR. Euro Surveill. 2020 Jan;25(3):2000045. doi: 10.2807/1560-7917.ES.2020.25.3.2000045. PubMed PMID: 31992387; PubMed Central PMCID: PMC6988269.

5. Josh Quick 2020. nCoV-2019 sequencing protocol v3 (LoCost). protocols.io https://protocols.io/view/ncov-2019-sequencing-protocol-v3-locost-bh42j8ye

6. Katoh K, Rozewicki J, Yamada KD. MAFFT online service: multiple sequence alignment, interactive sequence choice and visualization. Brief Bioinform. 2019 Jul 19;20(4):1160–1166. doi: 10.1093/bib/bbx108. PubMed PMID: 28968734; PubMed Central PMCID: PMC6781576.

7. Suchard MA, Lemey P, Baele G, Ayres DL, Drummond AJ, Rambaut A. Bayesian phylogenetic and phylodynamic data integration using BEAST 1.10. Virus Evol. 2018 Jun 8;4(1):vey016. doi: 10.1093/ve/vey016. PubMed PMID: 29942656; PubMed Central PMCID: PMC6007674.

8. Rambaut A, Holmes EC, O’Toole Á, Hill V, McCrone JT, Ruis C, et al. A dynamic nomenclature proposal for SARS-CoV-2 lineages to assist genomic epidemiology. Nat Microbiol. 2020 Nov;5(11):1403–1407. doi: 10.1038/s41564-020-0770-5. Epub 2020 Jul 15. PubMed PMID: 32669681.

